# Mental Health and Stress Level of Ukrainians Seeking Psychological Help Online

**DOI:** 10.1101/2022.08.19.22278660

**Authors:** Anastasiya-Mariya Asanov Noha, Igor Asanov, Guido Buenstorf

## Abstract

We screen the mental well-being and psychological distress of 1165 refugees, migrants, internally displaced, and non-displaced people from Ukraine who seek psychological help online in Ukraine and across 24 countries of the European Union. We see that more than half of the respondents exhibit low levels of mental well-being and high psychological distress, with 81% being at risk of depression and 57% having severe psychological distress. Refugees and internally displaced people in our sample show a particularly high risk of depression and severe psychological distress. Nevertheless, the majority of Ukrainians seeking psychological help still work, study, or volunteer, and those who do have relatively alleviated mental levels of well-being.

## Introduction

Due to the invasion of Ukraine, more than 9.2 million Ukrainians had to leave their home country, and more than 6.2 million Ukrainians remain internally displaced, giving rise to “the largest human displacement crisis in the world today” [1]. Moreover, these mass movements of people drastically differ from previously experienced in Europe [2], posing a challenge for quick (mental) health assistance and rapid (mental) health needs assessment [3]. Robust quantitative analysis is needed to understand the (mental) health conditions of the conflict-affected population [3]. The need for a uniform assessment is particularly pressing given that the Ukrainian mental health situation was a blind spot even before the invasion [4,5].

Aiming to address the need for robust quantitative evidence on the mental health conditions of Ukrainians seeking help, we provide information about the mental health status of refugees, migrants, internally displaced, and non-displaced people from Ukraine seeking psychological help. As part of the “Self-Help Online” program [6], we quickly screen 1165 program applicants in a uniform online survey for their mental health status and stress levels in Ukraine and across 24 countries in European Union. In addition, we elicit information about socio-demographic characteristics, current living conditions, as well as current and past location.

We find that the large majority of Ukrainians seeking help exhibit depressive syndromes and high levels of stress with probable serious mental illness that are considerable even for the population of refugees seeking help [7,8]. We observe that 81% of respondents are below the WHO-5 depression cutoff; 64% are below the MHI-5 severe depression cutoff; 57% of them are at risk of serious mental illness. We observe that respondents who are externally or internally displaced, respondents born in the Eastern part of Ukraine, and unemployed respondents exhibit particularly high levels of depressive syndromes and stress. However, even among the Ukrainians seeking help, we see that more than 40% of participants engage in volunteering activities. Those who do have relatively alleviated mental health conditions.

We collect and analyze unique and conceptually novel data to provide a systematic assessment of the mental health of refugees, migrants, internally displaced, and non-displaced people from Ukraine seeking help. Our data extends across 25 countries in a relatively large sample [7,8]. The uniform assessment of mental health status in our survey allows descriptive comparisons by migration status and other characteristics. To date, the United Nations High Commissioner for Refugees provided a survey of the profile and intentions of Ukrainian refugees in six European Union countries [9] and an online survey provides estimates of the labor market prospects of Ukrainian refugees in Germany [10]. Results of a recent meta-analysis of the mental health of forced migrants (refugees and internally displaced individuals) urge to assess mental health needs right after resettlement [11].

We also provide validation in the Ukrainian context of much-needed non-invasive tools required for screening mental health [4,5]. The existence of our data provides evidence of the methodological possibility to systematically identify, and reach out to, war-conflict-affected people seeking help across borders for telemental and online help [12].

## Results

Conflict-affected populations are likely to suffer from reduced mental well-being and high levels of stress. To understand the well-being of Ukrainians seeking help and the heterogeneity of mental health among them, we report descriptive results for the aggregate sample and by sub-groups. We focus on the following indicators of mental health: Mental well-being and probable depression based on the WHO-5 scale [13], probable severe depression and whether a person reported being happy based on MHI-5[14], as well as absolute stress and probable severe mental illness based on Kessler-6 (K6) [15].

### General pattern

The general pattern emerging from our data is worrying (see the first row of Table 1). Average respondents have low mental well-being, and most of them can be classified as having probable depression and a high level of stress. On average, respondents scored 37.46 points on the WHO-5 measure. To benchmark this value, consider that the Danish general population’s average score on the WHO-5 scale is 70 points [16], refugees and asylum seekers with psychological distress in cross-country study in Western Europe on average score ∼47 points [17], and a value ≤ 50 points is used to assign ‘screening diagnoses’ of depression [13]. We see that 81% of respondents are below the 50-point depression cut-off. Moreover, we see that 64% of respondents score below the severe depression cutoff based on MHI-5, whereas only 7% can be classified as happy. Finally, the average respondents score 13.15 on the K6 which measures psychological distress. 57% of respondents score 13 or higher on K6, indicating a high probability of having a serious mental disorder [14,15].

**Table 1:**
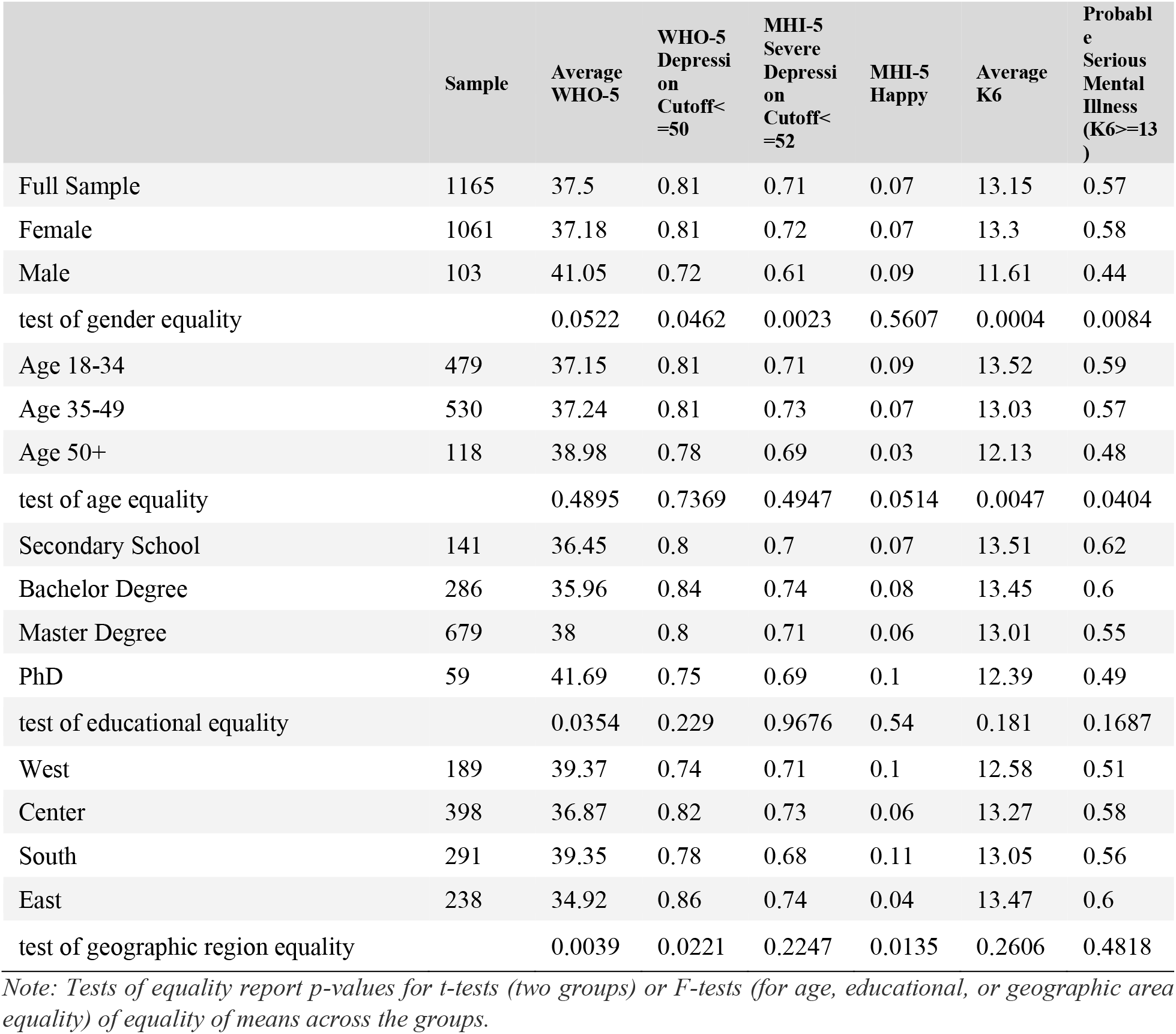
Mental health by socio-demographic characteristics

### Mental health by socio-demographic characteristics

Table 1 shows the breakdown of the mental health status by socio-demographic characteristics. 91.15% of respondents in our sample are women, which is expected given the gender characteristics of the displaced population and conflict-affected conditions of non-displaced individuals. We see in our sample that compared to men, women show somewhat lower mental well-being based on the WHO-5 scale and higher levels of depression. We also see a strong indication of higher psychological distress on K6 in women compared to men. Women score 13.3, whereas men score 11.16 on K6 (2.14 points difference). In turn, as the average woman scores 13.3, most of the women (58%) score above 13 indicating probable serious mental illness.

The age of respondents (based on the date of birth) ranges from 18 to 74. We see a homogenous pattern of low mental well-being and high depression levels among all age groups in our sample. However, we see a higher level of psychological distress in K6 among the younger cohort of respondents. The youngest cohort (18-34 years old) score 0.49 points more compared to the middle-aged cohort (35-49 years old), whereas the middle-aged cohort scored 0.9 points more than the oldest cohort (50+ years old). Higher psychological distress among the younger population is commonly observed in the general population even if one accounts for item bias [18].

About 87% of the respondents have at least a bachelor’s degree, which corresponds to 82.7% totally enrolled or 88.8% of women enrolled in tertiary education in Ukraine [19]. We see some level of heterogeneity by level of education in mental health well-being measured on the WHO-5 scale, but not in the level of depression or psychological distress.

Next, we break down the mental health status by macro-regions where the respondents were born: East (Donetsk, Kharkiv, Luhansk oblast’), South (Odesa, Mykolaiv Oblast, etc.), Center (Kyiv, Kyiv oblast, etc.), and West (L’viv, Volyn, Zakarpatska Oblast, etc.). Compared to respondents born in the West of Ukraine, individuals born in the East of the country show a lower level of mental well-being and a higher level of depression. Respondents born in the East score 4.45 points (11 percentage points) lower on the WHO-5 scale than people born in the West. ‘Screening diagnoses’ of depression are 12% more common among respondents born in the East than those born in the West (86% score below the depression cutoff in the East, whereas 74% of respondents score below the depression cutoff in the West). Only 4% of the respondents born in the East declare to be happy, which is less than half the (already low) value in the West, where 10% of respondents declare to be happy.

### Mental health by current migration status and (prior) region of residence

Fig. 1 illustrates the distribution of the mental well-being of Ukrainians seeking help based on the WHO-5 scale by their residence region before 2022 for the whole sample (Panel A) as well as by their current status of migration: non-displaced (Panel B), externally displaced (Panel C) and internally displaced (Panel D). Colors correspond to the average mental well-being on the WHO-5 scale, with darker red tones denoting lower mental well-being. We keep neutral yellow color in the residence region before 2022 if we have less than five Ukrainians seeking help from this region in our sample, to account for the differential spatial distribution of Ukrainians seeking help.

**Fig.1.**
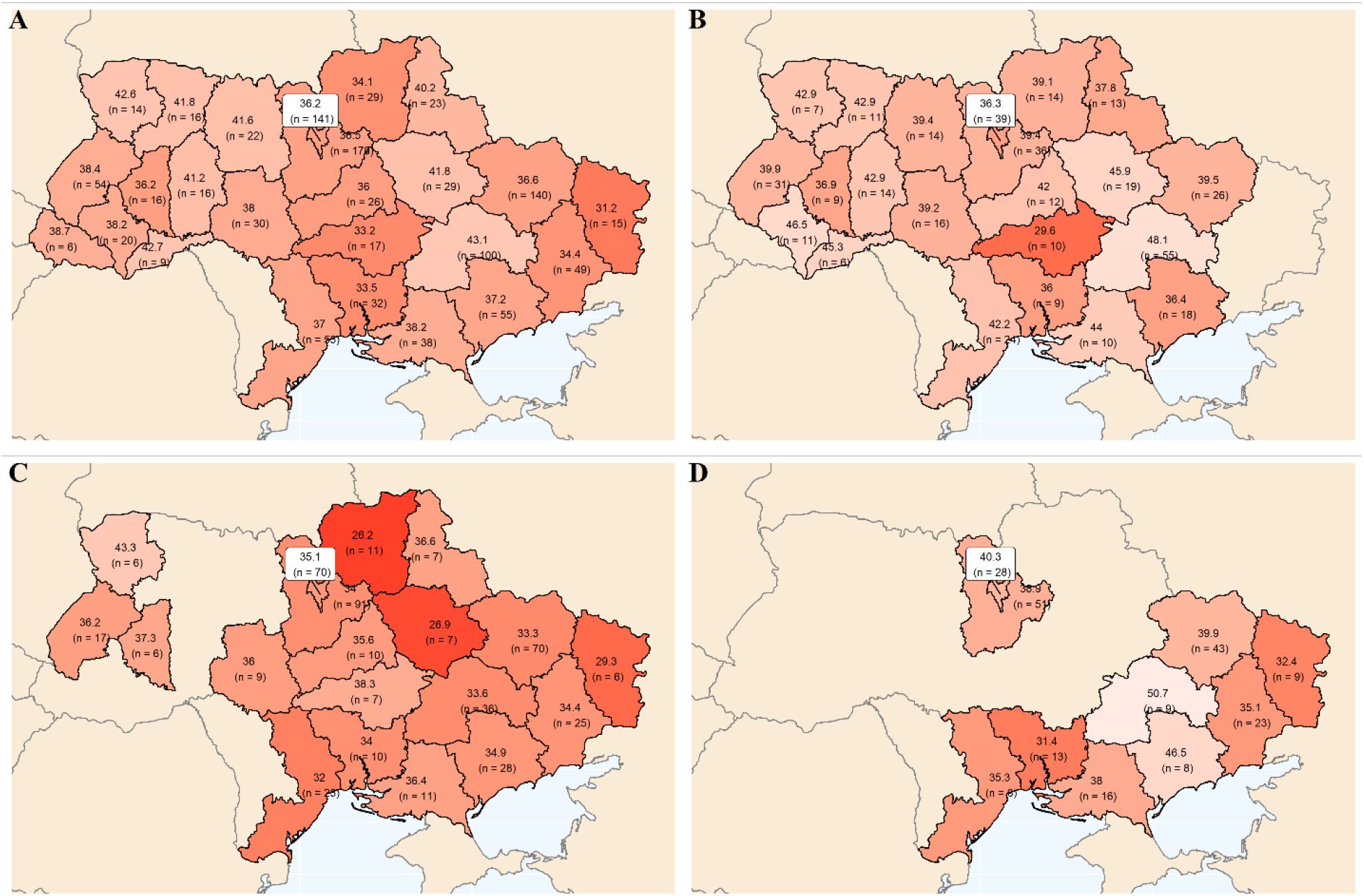
The distribution of the mental well-being of Ukrainians seeking help based on the WHO-5 scale by their residence region before 2022 for the whole sample (Panel A) for non-displaced (Panel B), externally displace (Panel C), internally displaced (Panel D). Darker red tones on map indicate lower mental well-being. Regions with less than 5 respondents are excluded.

We see in the top-left panel A of Fig. 1 that Ukrainians seeking help in our sample have low mental well-being on the WHO-5 scale, with an average not higher than 50 indicating “screening depression”. Darker red tones are more common in the Central and Eastern parts of the country. The top-right panel B of Fig. 1 depicts the mental well-being of non-displaced Ukrainians residing within the country, with a relatively homogenous pattern of mental well-being (with no respondents who currently reside in the Luhansk region and one respondent in the Donetsk region - not shown on the map). The bottom-left panel C of Fig. 1 shows the mental well-being of the externally displaced (refugees). Visual inspection of panels A to C of Fig. 1 suggests lower mental well-being of refugees compared to the whole sample of non-displaced people. Finally, the bottom-right panel D of Fig. 1 shows that internally displaced people seeking help were residing mostly in the South-eastern part of the country and Kyiv (city and oblast) before 2022.

Next, Fig. 2 shows density plots and boxplots of the distribution of psychological distress based on the K6 scale and grouped by current migration status. We see in Panel A of Fig. 2 that, while all groups show high levels of psychological distress, the distributions of psychological distress for the internally (yellow area) or externally (red area) displaced are located to the right of the distribution of non-displaced Ukrainians (blue area). This indicates that internally or externally displaced people show higher levels of psychological distress compared to the non-displaced. A higher level of psychological distress is especially pronounced for the externally displaced (refugees), where one can see that most of the refugees (a larger part of the distribution) show psychological distress above the 13-point cut-off of probable serious mental illness.

**Fig. 2.**
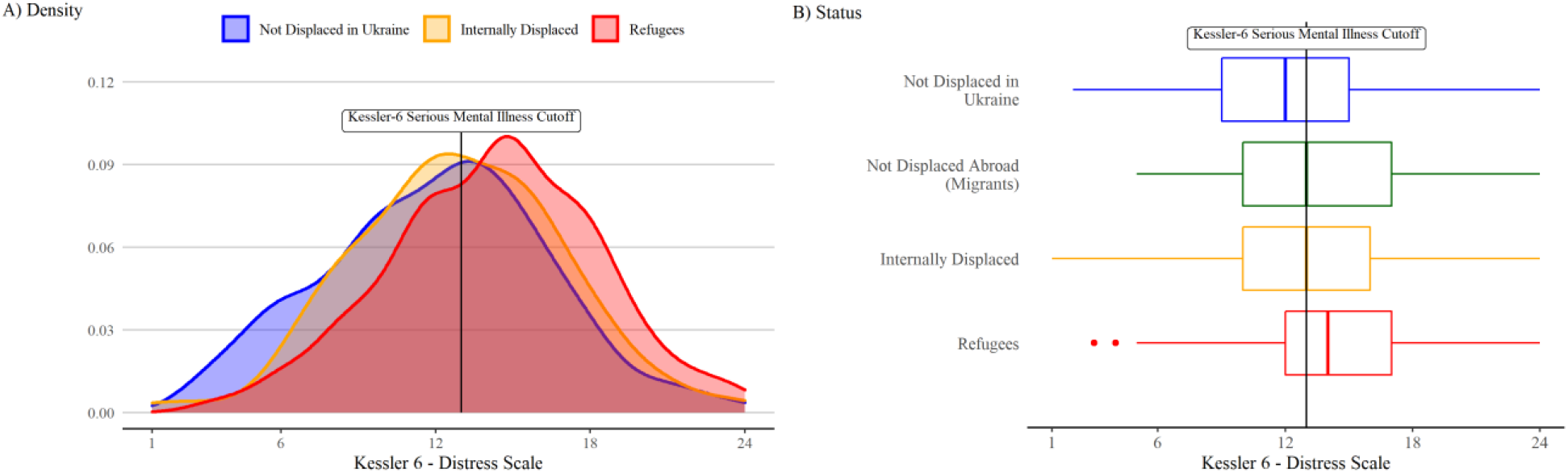
Density plots (Panel A) and boxplots (Panel B) of psychological distress based on the K6 scale grouped by current migration status. Migrants not reported on the density plot for clarity of presentation.

Panel B of Fig. 2 shows that the median externally displaced Ukrainian seeking help (vertical line in the middle of the box) has higher psychological distress than the median migrant, internally displaced, or non-displaced Ukrainian. The median non-displaced Ukrainian exhibits a high level of stress, which however is below the severe mental illness cut-off. The median migrant and the median internally displaced Ukrainian show psychological distress exactly at the level of the severe mental illness cut-off. However, median refugee psychological distress is above the severe mental illness cut-off.

Table 2 reports the breakdown of the mental health status by current migration status and other characteristics of Ukrainians seeking help. In line with a visual inspection, we observe the considerable gap between non-displaced people and refugees on all mental health indicators. Refugees score 6.71 points lower than non-displaced people on the WHO-5 scale. Probable depression on the WHO-5 scale is 16 percentage points more common among refugees seeking help than among non-displaced people (89% of refugees seeking help score below the depression cutoff, whereas 73% of non-displaced people seeking help score below the depression cutoff).

**Table 2:** Mental health by migration status and socio-economic characteristics

|  | Sample | Average WHO-5 | WHO-5 Depression Cutoff<= 50 | MHI-5 Severe Depression Cutoff<= 52 | MHI-5 Happy | Average K6 | Probable Serious Mental Illness (K6>=13) |
| --- | --- | --- | --- | --- | --- | --- | --- |
| Full Sample | 1165 | 37.5 | 0.81 | 0.71 | 0.07 | 13.15 | 0.57 |
| Not Displaced in Ukraine | 409 | 40.98 | 0.73 | 0.65 | 0.1 | 12.03 | 0.48 |
| Internally Displaced | 239 | 38.66 | 0.77 | 0.69 | 0.04 | 12.91 | 0.54 |
| Not Displaced, Abroad (Migrants) | 48 | 33.67 | 0.81 | 0.75 | 0.08 | 13.12 | 0.56 |
| Refugees | 469 | 34.27 | 0.89 | 0.78 | 0.06 | 14.25 | 0.66 |
| test of status equality |  | 0 | 0 | 0 | 0.0136 | 0 | 0 |
| Employed Full Time | 423 | 40.02 | 0.74 | 0.67 | 0.09 | 12.44 | 0.5 |
| Employed Part Time | 254 | 38.41 | 0.85 | 0.77 | 0.06 | 13.2 | 0.56 |
| Self-Employed | 53 | 40.68 | 0.75 | 0.6 | 0.11 | 11.74 | 0.51 |
| Unemployed | 276 | 33.59 | 0.88 | 0.74 | 0.04 | 14.05 | 0.67 |
| Student | 87 | 38.67 | 0.74 | 0.71 | 0.1 | 13.28 | 0.59 |
| Other | 72 | 30.72 | 0.89 | 0.75 | 0.04 | 14.53 | 0.67 |
| test of employment status equality |  | 0 | 0 | 0.0134 | 0.1137 | 0 | 0.0002 |
| Registered * | 592 | 35.78 | 0.85 | 0.75 | 0.06 | 13.78 | 0.61 |
| Not Registered * | 125 | 36.13 | 0.82 | 0.73 | 0.05 | 13.7 | 0.62 |
| test of registration status equality |  | 0.8254 | 0.3057 | 0.614 | 0.6606 | 0.8362 | 0.8214 |
| Receive Financial Support * | 386 | 34.89 | 0.87 | 0.75 | 0.05 | 13.87 | 0.62 |
| No Financial Support, Aware of Opportunities * | 169 | 39.36 | 0.79 | 0.7 | 0.06 | 13.12 | 0.56 |
| No Financial Support, Not Aware of Opportunities * | 154 | 33.87 | 0.86 | 0.81 | 0.04 | 14.36 | 0.68 |
| test of financial support equality |  | 0.0013 | 0.0543 | 0.2288 | 0.6884 | 0.0234 | 0.0865 |
| Studying | 713 | 38.75 | 0.77 | 0.68 | 0.08 | 12.87 | 0.55 |
| Not Studying | 452 | 35.52 | 0.86 | 0.76 | 0.05 | 13.6 | 0.6 |
| test of studying equality |  | 0.0004 | 0.0003 | 0.0681 | 0.0236 | 0.0047 | 0.0994 |
| Volunteering | 506 | 40.32 | 0.75 | 0.67 | 0.09 | 12.6 | 0.51 |
| Not Volunteering | 659 | 35.34 | 0.85 | 0.75 | 0.05 | 13.57 | 0.61 |
| test of volunteering equality |  | 0 | 0 | 0.0395 | 0.008 | 0.0001 | 0.0005 |

*Note: \* only present for displaced (externally and internally) participants. Tests of equality report p-values for t-tests (two groups) or F-tests (for migration status, educational, employment, or financial support equality) of equality of means across the groups.*

Moreover, we see that refugees score 2.25 points higher than non-displaced people on psychological distress (14.25 for refugees vs. 12.03 for non-displaced people in our sample). To benchmark this value, consider that the average score on the K6 scale in the US general population did not exceed 2.8 in the period from 1997-2007 [20], and the difference on the K6 scale between respondents in our sample born in the East and West is 0.89. In turn, probable serious mental illness measured on the K6 scale is 18 percentage points more prevalent among refugees seeking help than among non-displaced people (66% of refugees seeking help show indication of probable serious mental illness, whereas 48% of non-displaced people seeking help are at risk of probable serious mental illness).

As a robustness check of the descriptive disparity by current migration status, we assess this disparity by accounting for the time when respondents registered as users on the online platform through which the “Self-Help Online” program was provided (see Methods section for details). The time of registration is likely to correlate with the program and survey advertisement strategy. We run two simple linear regressions to assess the association between well-being on the WHO-5 scale or the psychological distress K6 scale and migration status including a dummy variable denoting whether users registered on the platform during the advertisement campaign and, thus, are likely to be brought by our external media campaign rather than the platform itself. The estimate and statistical significance of the refugee status barely change, indicating the robustness of the disparity between non-displaced individuals and refugees. As a further robustness check, we also use a machine learning approach to find and account for the potential confounders among a large set of controls with help of the double-lasso procedure [21]. The machine learning procedure selects a large set of controls (new user on the platform, East macro-region birthplace, etc.), yet the significant mental health penalty of the refugee status remains robust to the inclusion of these controls. These regressions are reported in Supplementary Table S1.

### Mental health by socio-economic status

We examine mental health status by employment and educational status (see Table 2). 7.5% of Ukrainians seeking help have student status in Ukraine or abroad, and 58% of Ukrainians seeking help in Ukraine or abroad have at least part-time employment, whereas 23% are unemployed. We see a strong heterogeneity in mental health status by employment and educational status across all indicators. 88% of unemployed Ukrainians seeking help are at risk of depression on the WHO-5 scale and score 14.53 on the K6 scale on average, whereas 74% of full-time employed Ukrainians are above the probable depression cut-off and score 12.44 on the K6 scale.

Our sample comprises non-negligible numbers of refugees and internally displaced people who did not register their residence and, thus, are not necessarily present in the official statistics. In our sample, 17% of refugees and internally displaced people did not register their residence (see Table 2). We, however, do not observe substantial differences in mental health between those who register and not. Moreover, we do not see a difference in mental health by registration status among refugees or internally displaced individuals (not reported in the table).

European Union countries provide financial support for refugees. Many refugees in our sample are aware of these opportunities and receive governmental financial support (social welfare, reimbursement for rent, etc.). 54% of refugees in our sample receive some financial support. 78% are aware of these opportunities, but the remaining 22% do not know about these opportunities and do not receive financial support. These 22% of refugees in our sample have lower mental well-being on the WHO-5 scale and higher psychological distress on the K6 scale than those who are aware of financial opportunities. Those who are not aware of financial opportunities (and thus stay without financial support) score 5.49 points lower on the mental well-being WHO-5 scale and 1.24 points higher on psychological distress than those who are aware of financial opportunities but do not receive financial support (relying on their own funds). 68% of those who are not aware of financial opportunities are above the cut-off for probable serious mental illness on the K6 scale.

Despite the circumstances, 61% of the Ukrainians seeking help engage in some form of studying (university, technical academy, online course, vocational training, etc.). Those who study (and are capable to study) report better mental well-being on all indicators compared to those who do not. Moreover, 43% of Ukrainians seeking help engage in volunteering activities themselves. Those who are (and are capable of) volunteering have relatively alleviated mental-health conditions compared to those who do not on all mental health indicators. Those who volunteer declare to be happy more than twice as often as those who do not (10% happy among volunteers, with only 4% happy among those who do not).

### Mental health of displaced by current location

We have a look at the mental health status among the internally or externally displaced (refugees) by their current country of location. Panel A of Fig. 3 illustrates the average mental well-being of Ukrainians seeking help based on the WHO-5 scale by country of location (darker red tones indicate lower mental well-being). Visual inspection shows a relatively homogenous pattern of low mental well-being among displaced Ukrainians seeking help, with a slightly better situation among displaced Ukrainian who stayed in their home country. Panel B of Fig. 3 shows that the median displaced Ukrainian seeking help (line in the middle of the box) scores below 50 on the WHO-5 scale in any country indicating that “screening depression” is prevalent irrespective of the country of location.

**Fig. 3.**
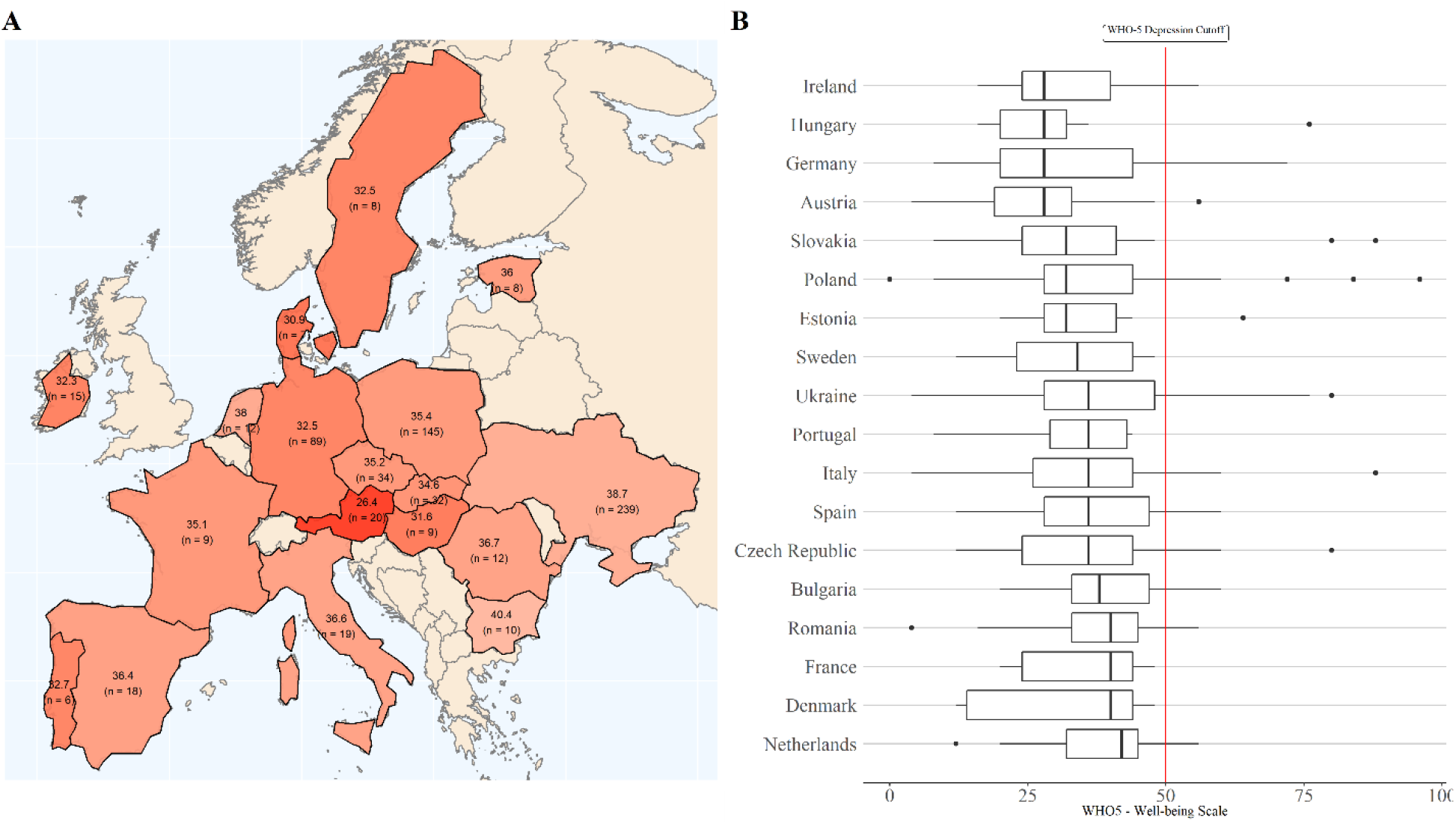
Distribution of average (Panel A) and boxplots (Panel B) of mental well-being of Ukrainians seeking help based on the WHO-5 scale by country of location. Darker red tones on the map indicate lower mental well-being. Countries with less than 5 respondents are excluded.

## Discussion

We screen refugees, migrants, internally displaced and non-displaced people from Ukraine seeking help in Ukraine and 24 European countries for their mental health. Most Ukrainians seeking help suffer from low mental well-being and high stress levels indicating they are at risk of depression and severe psychological distress. Refugees and internally displaced individuals in our sample show a particularly high risk of depression and severe psychological distress. Those who are unemployed and those who do not know about governmental support programs show especially worrying levels of mental health status. Nevertheless, we see that the majority of Ukrainians seeking help do work, study, or volunteer. Those who do have relatively alleviated mental well-being levels.

Our results indicate that despite considerable efforts to improve the mental well-being of Ukrainians [9], there is an urgent need to develop methods to improve the mental health situation among the conflict-affected population at scale. With an online platform, we were able to reach out to a large number of Ukrainians seeking help in a short time period, across 25 countries and irrespective of their current status. This shows the potential of complementing existing offline psychological services with online tools that can be provided to anyone who has access to the Internet. Moreover, programs that inform about available support options or help to improve the labor situation among the conflict-affected population potentially can be bundled with mental health interventions to achieve better results. Finally, while we screen the people who were seeking help as part of the “Self-Help Online” program, systematic screening for the mental health of the conflict-affected population can help to reach out to those who are in need, thus augmenting targeted mental health policies.

## Methods

### Context

We recruit and survey participants in the study as a part of our “Self-Help Online” program for adult Ukrainians [6]. Recruitment, data collection procedure, and study were approved by the central ethical committee of the University of Kassel (decision letter zEK-033 from 25.05.22). After providing an information sheet that explains the details of the study and the program in plain language, we survey online participants who actively consent to data collection for scientific purposes and participation in the program. Our “Self-Help Online” program provides all participants with extensive online information about psychological (and other) support offered online to Ukrainians in Ukraine and other countries and tests the “Self-Help Online” course built on the WHO Self-Help+ offline materials recommended for online adaptation by WHO [22]. The program and surveys were implemented in the Ukrainian language, but we offered respondents to get the materials in other languages.

### Sample

We recruited participants within a compressed two-week timeframe (from 22.06.2022 to 06.07.2022) to deliver the materials quickly, increasing the frequency of program advertising closer to the deadline to minimize participants’ waiting time. We used two main strategies in recruiting participants: (a) a social media campaign and (b) internal resources of the Ukrainian online educational platform, where we deliver the program.

For the social media campaign, first, we provided two-day A/B testing of the marketing materials (texts, visuals, etc.) on the targeted group on Facebook and Instagram (adult Ukrainians, and adult Ukrainians who are currently outside their country based on Meta geolocation). Second, we set up advertisements on Facebook and Instagram, budgeting advertisements to Ukrainian and Russian-speaking users across three dimensions: (1) current geolocation; (2) gender, and (3) time before the program. We budgeted spending per European Union in the proportion of refugees in the respective country out of the total number of Ukrainian refugees in European countries, with 90% of the budget allocated to females and 66% of the budget spent one week before the start of the program. This part of the advertisement campaign reached 1.19 million people at least once. Five days before the registration, we posted advertisements manually on Facebook and Telegram groups for Ukrainian refugees (total number of members of these groups: 1.9 million people). The Ukrainian online educational platform advertised the course to their users by putting the program on their main page, and mailing it to their users (at first, only our program and right before the end of registration together with other online psychological courses).

These efforts resulted in 1 201 registrations with effective surveys within two weeks, out of which 35 were located neither in Ukraine nor the European Union, and one double registration. Accordingly, our survey sample consists of 1 165 participants. Though our sample should by no means be considered representative of the population, but only of those seeking help online (see discussion of non-random survey bias in [23]), we see that generally proportions of participants by country correspond to the proportions of refugees in these countries and that proportions of participants by birth region (oblast’) correspond to the proportions of the population in these regions in Ukraine. We naturally have mostly female participants, with 104 male participant, and younger participants on average (Median Age: 36; Minimum Age: 18; Maximum Age: 74: SD=10.11) than the population average of 41.2. About 87% have at least a bachelor’s degree, which corresponds to 82.7% totally enrolled or 88.8% of females enrolled in tertiary education, as Ukraine is among the most highly educated societies in the world [19].

### Tools: Mental Health Scales

To screen for mental health conditions, we used widely accepted short (not more than six items) non-invasive tools with high construct validity: WHO-5[13], MHI-5[14], and K6[15] scales. These tools allowed us to minimize screening time and avoid potential re-traumatization while assessing respondents’ mental well-being and psychological distress.

We also provide validation in the Ukrainian context of much-needed non-invasive tools to screen for mental health conditions [4,5]. The WHO-5[13], MHI-5[14], and K6[15] scales were translated from English to Ukrainian by a leading researcher and then back-translated by a research assistant. After research team deliberation based on a third opinion, rare discrepancies between translation and back-translation were harmonized. The survey was implemented on the online platform aiming as closely as possible to resemble the visual representation of the original scales.

The timeframe of mental health status was formulated as in the original scales: We asked participants about their mental health status over the last two weeks in the case of WHO-5, last month in the case of MHI-5, and 30 days in the case of the K6 scale. Respondents were asked to rate how often they experience a certain state in each item of each scale. Then, in the case of the 5-item scale WHO-5, each item was scored from 5 (all of the time) to 0 (none of the time). The score from each item was summed up (ranging from 0 to 25) and multiplied by 4. In the case of the 5-item scale MHI-5, each item was scored from 1 (none of the time) to 6 (all of the time). The score from each item was summed up (ranging from 1 to 30) and the minimum possible score (5) was subtracted. As the last step, the score was divided by 25 (possible score range) and multiplied by 100. Participants who reported most or all of the time in the item “During the past month, how much of the time were you a happy person?” of the MHI-5 scale were classified as “Happy”. In the case of the 6-item scale K6, each item was scored from 4 (all of the time) to 0 (none of the time). The score from each item was summed up (ranging from 0 to 24).

All three scales – WHO-5, MHI-5, and K6 – show good internal consistency in our sample: Cronbach’s alpha values are 0.84 for the WHO-5 scale, 0.82 for the MHI-5 scale, and 0.83 for the K6 scale. The Pearson’s correlation coefficient between well-being scales WHO-5, and MHI-5 is 0.7, whereas between well-being and psychological distress scales WHO-5, and K6 is -0.65, or MHI-5 is -0.79. Scales in Ukrainian are available upon request.

### Tools: Socio-demographic characteristics and current status

We also elicit a set of socio-economic and demographic characteristics as well as the current status. Here we provide a description of the variables used to explore the heterogeneity of mental health among respondents in this study.

Gender was elicited with multiple answer questions: Female, Male, Other. We have one declared non-binary case in the sample. Age was calculated based on the date of birth of the respondent. In a few cases, the year of birth was set to 2022, or in one case to 1880, these values were set to NA.

Education level can take on the following categories: Secondary School (if the participant has completed basic secondary education (9 years) or full secondary education (11 years)), Bachelor’s degree if the participant has received a bachelor’s diploma (bakalavr), Master’s degree is the participant has received a Master’s degree (mahistr) and a Ph.D. if the participant is a Doctor of Sciences (Doktor Nauk).

We rely on the widely used classification of macro-regions by the Kyiv International Institute of Sociology reports [24]. The Western macro-region includes Volyn, Rivne, Lviv, Ivano-Frankivsk, Ternopil, Khmelnytskyi, Zakarpattia, and Chernivtsi oblasts; the Central macro-region includes Zhytomyr, Vinnytsia, Kirovohrad, Cherkasy, Poltava, Sumy, Chernihiv, Kyiv oblasts and the city of Kyiv; the Southern macro-region consists of Dnipropetrovsk, Odesa, Mykolayiv, Kherson, Zaporizhzhia oblasts and Crimea; the Eastern macro-region includes Kharkiv, Donetsk and Luhansk oblasts.

The classification of the current status of the respondent is based on two questions: “Did you have to leave your place of residence in 2022?” and “In which country are you currently located?” Respondents who did not have to leave their place of residence and who are currently located in Ukraine are considered to be “Not Displaced in Ukraine”. Respondents who had to leave their place of residence and are currently located in Ukraine are considered to be “Internally Displaced”. Respondents who had to leave their place of residence and are currently located in any EU country are considered to be “Refugees”. Finally, respondents who did not have to leave their place of residence and are currently located in any EU country are considered to be “Migrants”.

Employment or educational status includes full-time employed, part-time employed, self-employed, unemployed respondents, students, and “Other” for participants who answered “not employed and not looking for work” or “I don’t know”.

Registration status among displaced participants is determined by the question: “Did you manage to register your place of residence in the local government (city council, village council, etc.)?” Financial support status is determined by the question: “Are you currently receiving any social help from the local government (social/welfare transfers, reimbursement for rent, etc.)?” with the answer options: “Yes”, “No, but there is an opportunity for it” and “No, there is no opportunity to receive social help that I know of”.

Whether participants engage in any studying activities is determined by the question: “Are you currently enrolled in any form of education such as a university, technical academy, online course, vocational training, or other?” Volunteering status is determined by the question: “How many hours in the past week did you spend volunteering (doing any work for free)?” If the participant declares to spend at least one hour volunteering, we classify the participant as volunteering (43% of respondents in our sample). In our sample, the average respondents spend 3.67 hours volunteering, and the average respondents who volunteer spend 8.46 hours volunteering (values winsorized at 1% level).

## Data Availability

All data produced in the present study are available upon reasonable request to the authors

## Supplementary Materials

**Table S1:**
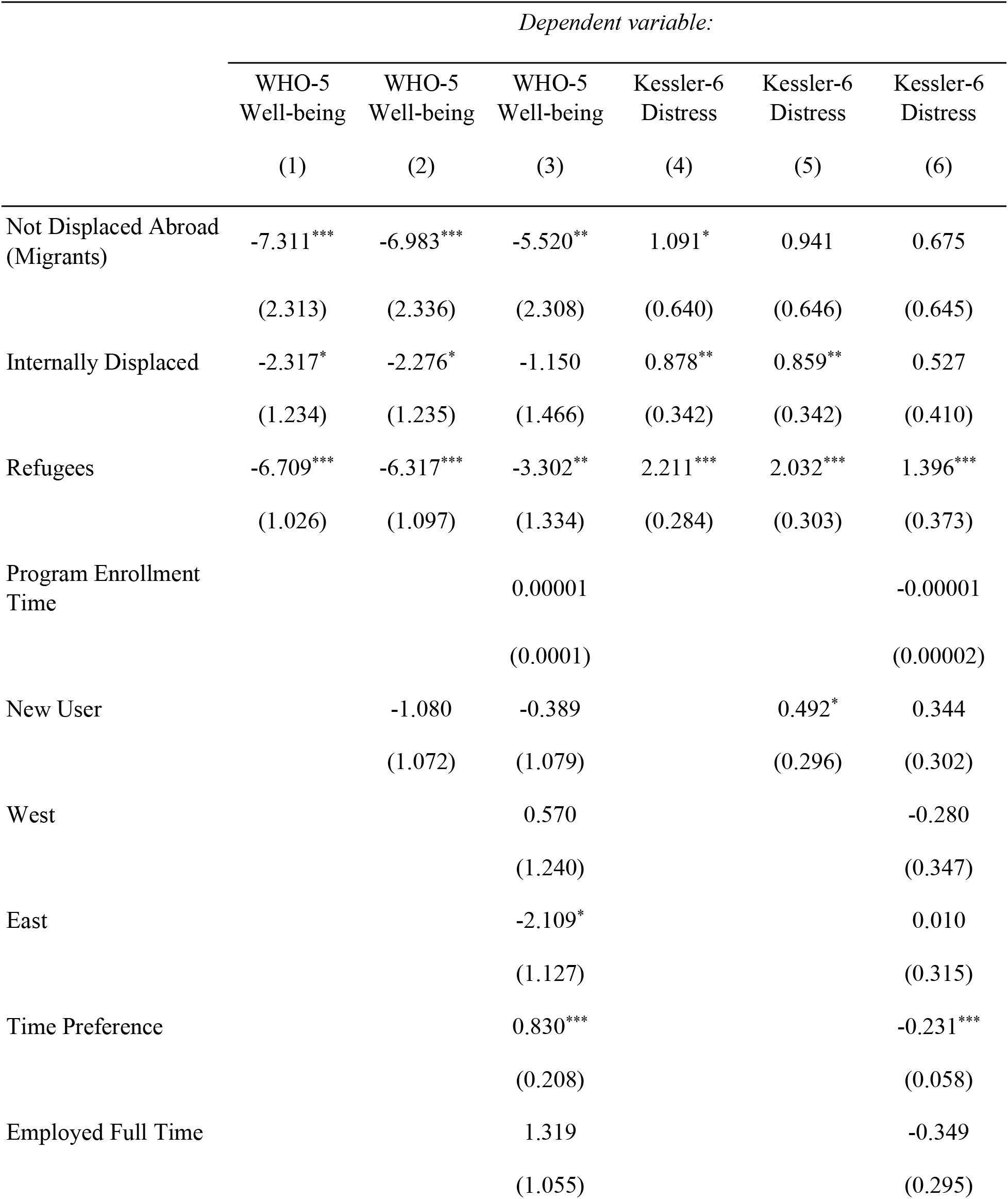
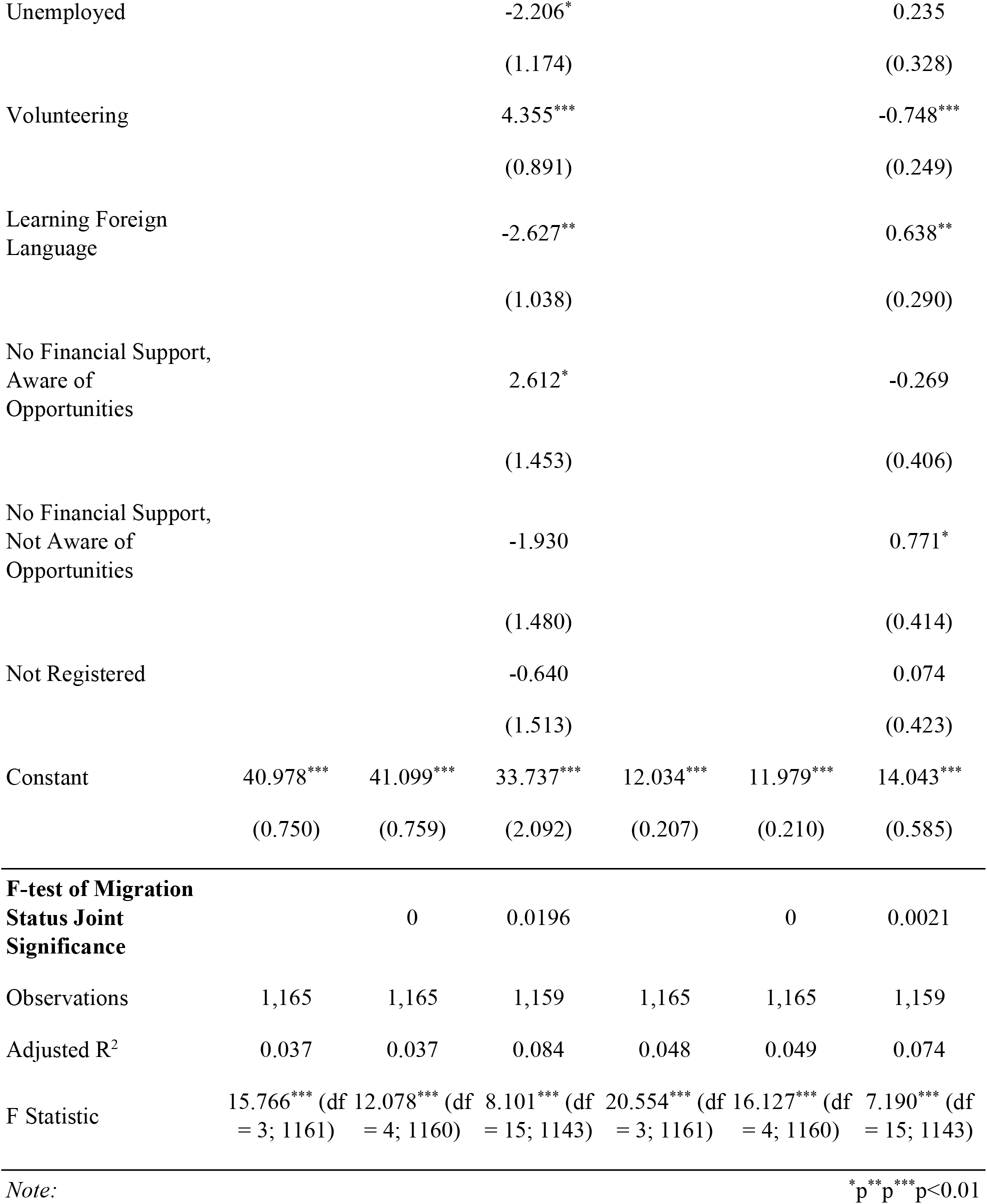
Robustness check of Health Status Disparity by Migration. (Reference group not displaced).

## Notes

We would like to thank INCHER-Kassel for support of the project. We are grateful to Thomas Kailer and Georg Krücken for kind support of the project from initial stage. We would like to thank Dariia Melnyk, Pia Schoch, Jana Zaremba for excellent research assistance and Alya, Iuliia Khodakivska, Andrii Parhomenko, Ivan Primachenko for technical support. Special thanks to Lesha and Sasha. We are also extremely grateful to Serhiy Kryzhnenko.

### Competing Interest Statement

The authors have declared no competing interest.

### Funding Statement

This study was funded by the University of Kassel International Center for Higher Education Research (Germany)

### Author Declarations

The Central Ethics Committee of the University of Kassel gave ethical approval for this work on 25/05/2022 (Approval letter ref: zEK-33). (Geschaftsstelle der zentralen Ethikkommission der Universitat Kassel Monchebergstr. 19 34125 Kassel, Germany;)

